# Causality between Peripheral Immune Cell Counts and Membranous Nephropathy: A Bidirectional Mendelian Randomization Study

**DOI:** 10.1101/2023.11.27.23299065

**Authors:** Zhihang Su, Liu Wen, Xiangning Feng, Qijun Wan

**Author notes:** **Correspondence:** Corresponding Author: Qijun Wan.

## Abstract

**Background:** Membranous nephropathy (MN), an autoimmune disease, has not yet been fully elucidated regarding its relationship with immune cells.

**Methods:** As a primary method in this Mendelian randomization (MR) analysis, we employed the Inverse Variance Weighted (IVW), Wald Ratio (WR), Steiger filtering, Weighted Median, Weighted mode, MR-Egger, Mendelian randomization pleiotropy residual sum, and outlier (MR-PRESSO) and Leave-one-out sensitivity test. Reverse MR analysis was utilized to investigate whether MN affects immune cells.

**Results:** After False Discovery Rate multiple correction (threshold was 10%), it was determined that CD45RA^+^ CD8^+^ T cell (IVW OR=1.074, 95%CI: 1.034-1.114, P=0.0001, P^FDR^=0.036), Effector Memory CD8^+^ T cell (IVW OR=0.929, 95%CI: 0.895-0.964, P=0.0001, P^FDR^=0.036), HLA DR^+^ CD8^+^ T cell (IVW OR=0.921, 95%CI: 0.884-0.960, P=0.0001, P^FDR^=0.036) are significantly affected by MN. CD28^+^ CD45RA^-^ CD8^+^ T cell (WR OR=0.513, 95%CI: 0.357-0.793, P=0.0003, P^FDR^=0.061), CD8 on Terminally Differentiated CD8^+^ T cell (WR OR=0.378, 95%CI: 0.219-0.652, P=0.0005, P^FDR^=0.061), and HVEM on Effector Memory CD4^+^ T cell (WR OR=0.378, 95%CI: 0.219-0.652, P=0.0005, P^FDR^=0.065), among others, are considered immune cells that possess protective factors against MN. However, HLA DR on CD33^+^ HLA DR^+^ CD14^-^ (WR OR=1.910, 95%CI: 1.269-2.876, P=0.002, P^FDR^=0.065), CD80 on granulocyte (WR OR=2.606, 95%CI: 1.417-4.792, P=0.002, P^FDR^=0.065), and CD62L^-^ plasmacytoid Dendritic Cell (WR OR=1.508, 95%CI: 1.138-1.998, P=0.004, P^FDR^=0.071) pose a risk to MN. When increased by one standard deviation in peripheral blood, these immune cells positively or negatively impact the risk of MN with an odds ratio (OR).

**Conclusions:** The MR analysis unveiled the association between immune cells and MN. We have deepened our understanding of the pathogenic mechanisms linking MN and immune cells and also identified new targets for immunotherapy in MN.

## 1 Introduction

Membranous nephropathy, an autoimmune disease, is characterized by the formation and deposition of immune complexes on the basement membrane, resulting in the thickening of glomerular capillary walls(van de Logt et al. 2019). This can lead to pathological and physiological disruptions of the podocyte structure, resulting in proteinuria. Some patients with membranous nephropathy experience mild symptoms. At the same time, severe cases may present with decreased serum albumin levels and generalized edema, known as nephrotic syndrome, and can progress to end-stage kidney disease. Approximately 80% of patients with membranous nephropathy (primary membranous nephropathy) have no identifiable cause(Couser 2017). In contrast, the remaining cases, known as secondary membranous nephropathy (SMN), are connected with other conditions such as lupus erythematosus, hepatitis B virus infection, and malignant growths. The main target antigens for primary membranous nephropathy are PLA2R, THSD7A, NELL1, SEMA3B, and PCDH7(Sethi 2021). While antigen-specific T cells and B cells have not yet been characterized in primary membranous nephropathy (PMN), autoantibodies targeting these antigens have been identified(Chung et al. 2022). Two-sample Mendelian randomization (MR) employs instrumental variables (IVs), which are proxy variables representing genetic variations, to simulate the causal impact of exposure on outcomes(Sekula et al. 2016). Typically, single nucleotide polymorphisms (SNPs) identified in genome-wide association studies (GWAS) are employed as genetic instruments to estimate the causal effect of exposure on outcomes(Sekula et al. 2016). Simultaneously, we aim to minimize the impact of confounding factors and reverse effects. Unlike traditional observational research, MR can mitigate the impact of confounding factors(Georgakis and Gill 2021). Previous studies have shown a correlation between membranous nephropathy and inflammation and the immune response(Ronco et al. 2021). Nevertheless, there is a lack of research analyzing the specific cell subpopulations associated with MN. Our study aims to establish the identity of the specific cell subpopulations connected with MN. The study design is depicted in Figure 1. Initially, we will employ MR using data from the Blood Cell Consortium(BCX) meta-analysis(Vuckovic et al. 2020), Peripheral Immune Cell GWAS summary statistics by Orr et al.(Orrù et al. 2020), and membranous nephropathy GWAS data from the Kiryluk Lab to identify the specific cell subpopulations associated with MN(Xie et al. 2020).

**Figure 1:**
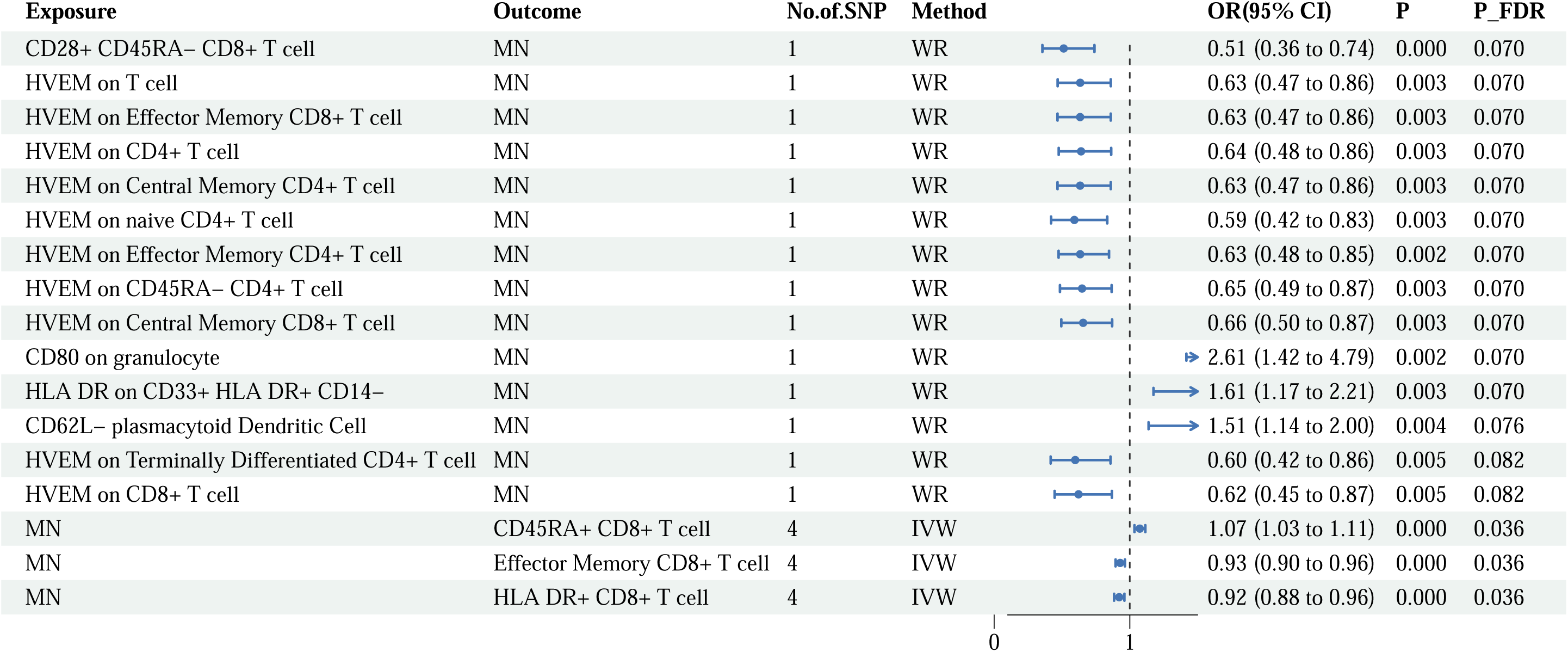
Flowchart for the Mendelian randomization analysis exploring effects of blood cell counts on membranous nephropathy. SNP, single nucleotide polymorphism; Leu, leukocyte; Lym, lymphocyte; Mono, monocyte; Neutro, neutrophil; Eosino, Eosinophil; Baso, basophil; IVW, Inverse variance weighted; MR-PRESSO, Mendelian Randomization Pleiotropy RESidual Sum and Outlier.

## 2 Methods and materials

### 2.1 Data of exposure

The effect estimates of single nucleotide polymorphisms (SNPs) linked to peripheral blood cell counts, such as total leukocytes, lymphocytes, monocytes, neutrophils, eosinophils, and basophils, are obtained from the meta-analysis conducted by the Blood Cell Consortium (BCX), comprising data from 563,085 individuals of European descent(Vuckovic et al. 2020).

To analyze lymphocyte subpopulations, which encompass T cells [naïve, central memory (CM), effector memory (EM), terminally differentiated (TD), regulatory (Treg), and natural killer (NKT) cells], as well as B cells (naïve, unswitched memory, switched memory, and transitional memory cells), we employed GWAS data obtained from Orr et al. This study examined 3,757 individuals from the Sardinian founder population using flow cytometry(Orrù et al. 2020).

### 2.2 Data of outcome

We have chosen Idiopathic Membranous Nephropathy (IMN) as the outcome variable. The data was obtained from a Genome-Wide Association Study (GWAS) focused on primary IMN, involving 2150 cases and 5829 controls across five European cohorts(Xie et al. 2020). All patients in the database diagnosed with membranous nephropathy were confirmed through kidney biopsy. Furthermore, the database excluded secondary factors such as drugs, malignant tumors, infections, or autoimmune diseases, ensuring the focus on IMN. The study population comprised individuals of European descent.

### 2.3 Mendelian Randomization

We conducted bidirectional MR to examine the causal connection between MN and peripheral blood cells as well as immune cell subpopulations. In cases where only one instrumental variable was available, we employed the Wald ratio (WR) as the primary method for Mendelian randomization (MR) analysis. In scenarios involving multiple instrumental variables, we utilized the inverse variance weighted (IVW) method as the main approach to estimate causal relationships. Additionally, we employed MR-Egger, Weight median, and Weighted mode as supplementary methods. Given the multiple analyses performed, there exists a potential for obtaining false positive results in Mendelian randomization (MR). We employed the false discovery rate (FDR) methodology for multiple tests to address this concern, setting the threshold at 10%(Van den Eynden et al. 2018; Kang et al. 2010; Bouras et al. 2022).

### 2.4 Selection of instrumental variables

Correlation: Our objective was to establish the individual associations of instrumental variables (IVs) with peripheral blood cells and lymphocyte subpopulations. We conducted a preliminary screening using a P-value threshold 5e-08, which confirmed the robust correlation between the instrumental variables and the aforementioned exposures. IVs with F < 10 [computed according to the F formula: F= R2(n - k - 1)/ k (1 - R2), F’= (β/SE)^2^] were excluded to strengthen the association between the instrumental variables and the exposures(Burgess, Thompson, and CRP CHD Genetics Collaboration 2011; de Klerk et al. 2023).

Independence: To guarantee the independence of variations in the instrumental variables (IVs), we conducted clustering using the linkage disequilibrium (LD) reference panel derived from the 1000 Genomes Project(“High-Coverage Whole-Genome Sequencing of the Expanded 1000 Genomes Project Cohort Including 602 Trios - PubMed” n.d.). At each locus, we retained the single nucleotide polymorphism (SNP) with the lowest p-value, excluding any other SNPs within a 10000 kb window with an R2 < 0.001 correlation with the IVs.

Exclusion criteria: We eliminated IVs with outcome p-values < 5e-05, as they could potentially be associated with MN. This step improved the accuracy of our MR analysis.

Confounding factors were identified using phenotype scanning. If a factor was found to influence both the exposure and the outcome simultaneously, it was excluded from subsequent analysis. We will exclude specific phenotypes, such as asthma, inflammatory bowel disease, and systemic lupus erythematosus, that can potentially impact peripheral blood immune cell counts.

The analysis encompassed a total of 790 instrumental variables.

### 2.5 Sensitivity analysis and directional test

After conducting the MR analysis, it is essential to validate the results through sensitivity analysis and directional tests. MR analysis is susceptible to issues such as horizontal pleiotropy, heterogeneity, and reverse causality. We employ the MR-Egger test and Mendelian randomization pleiotropy residual sum and outlier (MR-PRESSO) to evaluate the presence of horizontal pleiotropy in the instrumental variables. In order to examine heterogeneity in the instrumental variables, we utilize the IVW, MR-Egger, and Cochran’s Q test. For directional tests, we employ the Steiger filtering test and reverse MR analysis. The Steiger filtering test is employed to ensure the accuracy of the positive direction. In the reverse MR analysis, we interchange the exposure and outcome variables and conduct MR analysis again.

## 3 Results

Using bidirectional Mendelian randomization, we observed no causal relationship between peripheral blood cell counts, specifically lymphocyte count and MN (Figure 2). Furthermore, we did identify causal relationships between specific lymphocyte subpopulations and MN (Figure 3). In the analysis results of this subgroup, which specifically examined the causal relationship between lymphocyte subpopulations and MN, After False Discovery Rate multiple correction (threshold was 10%), it was determined that CD45RA^+^ CD8^+^ T cell (IVW OR=1.074, 95%CI: 1.034-1.114, P=0.0001, P^FDR^=0.036), Effector Memory CD8^+^ T cell (IVW OR=0.929, 95%CI: 0.895-0.964, P=0.0001, P^FDR^=0.036), HLA DR^+^ CD8^+^ T cell (IVW OR=0.921, 95%CI: 0.884-0.960, P=0.0002, P^FDR^=0.036) are significantly affected by MN. CD28^+^ CD45RA^-^ CD8^+^ T cell (WR OR=0.513, 95%CI: 0.357-0.793, P=0.0003, P^FDR^=0.070), HVEM on naive CD4+ T cell (WR OR=0.591, 95%CI: 0.420-0.833, P=0.0027, P^FDR^=0.070), and HVEM on CD8+ T cell (WR OR=0.622, 95%CI: 0.446-0.868, P=0.005, P^FDR^=0.082), among others, are considered immune cells that possess protective factors against MN. However, HLA DR on CD33^+^ HLA DR^+^ CD14^-^ (WR OR=1.910, 95%CI: 1.269-2.876, P=0.002, P^FDR^=0.070), CD80 on granulocyte (WR OR=2.606, 95%CI: 1.417-4.792, P=0.002, P^FDR^=0.070), and CD62L^-^ plasmacytoid Dendritic Cell (WR OR=1.508, 95%CI: 1.138-1.998, P=0.004, P^FDR^=0.076) pose a risk to MN. When increased by one standard deviation in peripheral blood, these immune cells have a positive or negative impact on the risk of MN with an odds ratio (OR) as indicated. This suggests that an increase in the counts of specific lymphocyte subpopulations may increase the risk of developing MN. Conversely, MN patients may exhibit elevated counts of certain lymphocyte subpopulations, leading to downstream effects that could be associated with diseases related to these subpopulations. The sensitivity analysis conducted on the aforementioned MR results indicates that horizontal pleiotropy and heterogeneity can be disregarded (p > 0.05).

**Figure 2:**
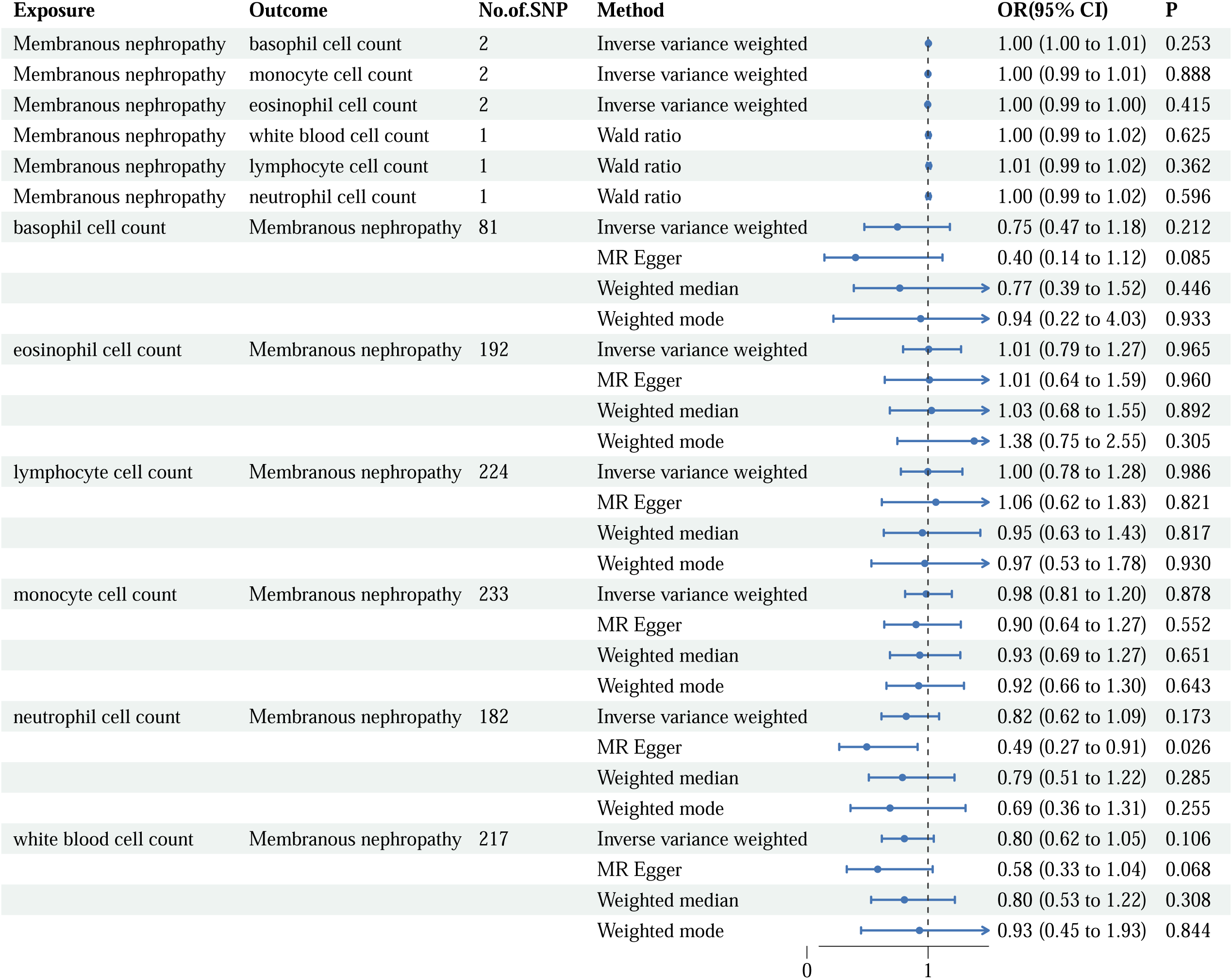
Mendelian randomization results of the association between blood cell counts and risk of membranous nephropathy. OR, odds ratio; CI, confidence interval.

**Figure 3:**
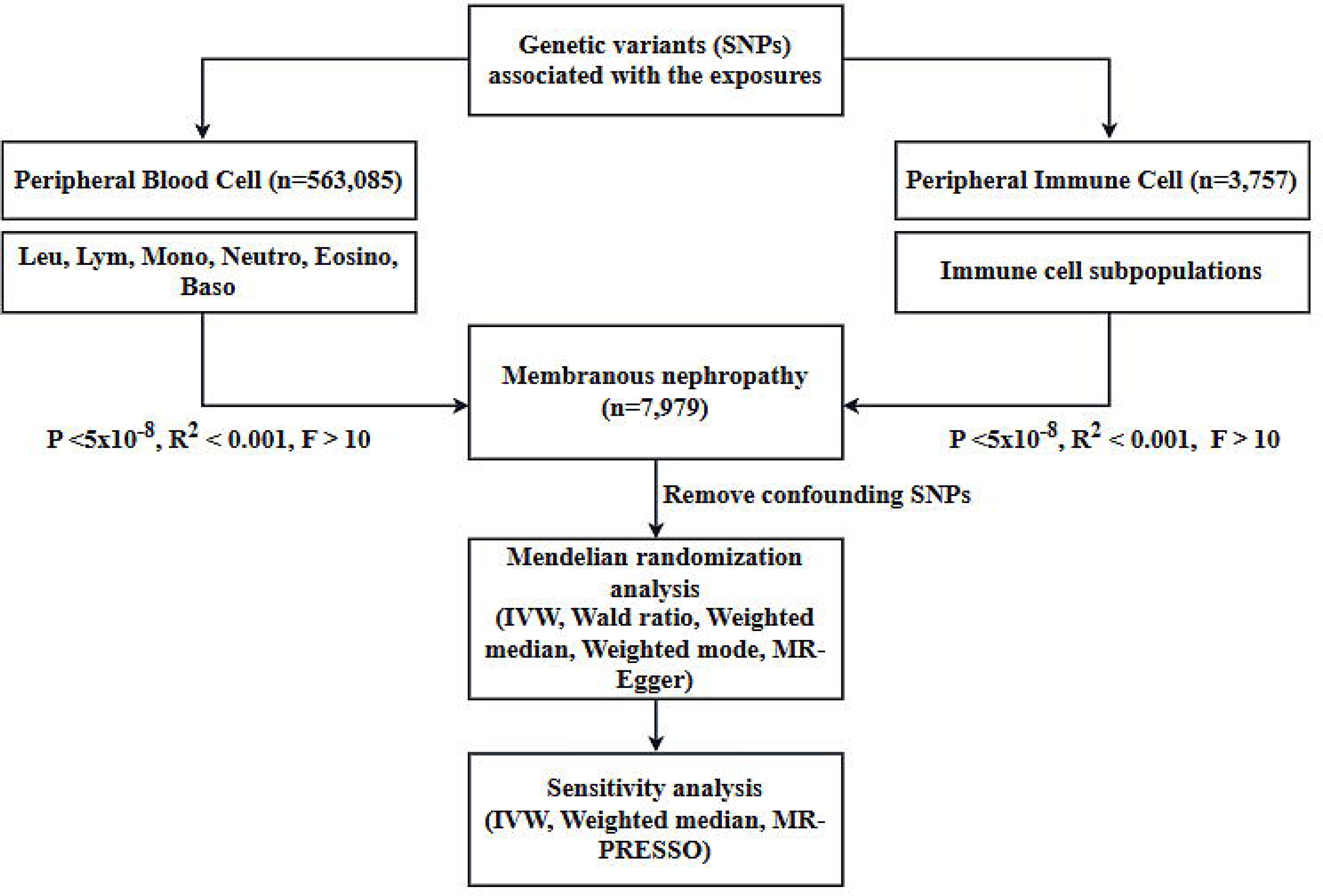
Mendelian randomization results of the association between immune cell counts and risk of membranous nephropathy. WR, Wald ratio; IVW, Inverse variance weighted; OR, odds ratio; CI, confidence interval; MN, membranous nephropathy; FDR, false discovery rate.

## 4 Discussion

Genome-wide association studies (GWAS) have recognized multiple reproducible genetic associations between common single nucleotide polymorphisms (SNPs) and both common autoimmune diseases and immune-mediated disorders(Cotsapas et al. 2011; Gutierrez-Arcelus, Rich, and Raychaudhuri 2016). Some of these associations are observed across multiple phenotypes. From a genetic standpoint, this confirms the interconnectedness of these immune-related diseases. In clinical practice, it is frequently observed that certain cases, like systemic lupus erythematosus, can result in secondary membranous nephropathy(Mok 2009). Similarly, patients with idiopathic membranous nephropathy may concurrently develop autoimmune diseases, including systemic lupus erythematosus and rheumatoid arthritis(Miyazaki et al. 2023; Caza et al. 2021). However, during MR analysis, we treat these phenotypes, which have the potential to cause changes in immune cell counts, as confounding factors. Eliminating these confounding factors prior to analysis can produce more accurate results.

Although distinct diseases necessitate varying dosages, there is often an overlap in the utilization of immunosuppressants and hormones for their treatment(Lazar and Kahlenberg 2023; Burmester and Pope 2017). Therefore, it is imperative to comprehend the impact of their respective targets on gene regulation and immune function. Immunophenotypes, including signaling responses, immune cell abundance, and serum cytokine levels, have the potential to determine pivotal immune actions associated with the risk of autoimmune disorders. Consequently, the identification of crucial immunophenotypes and the elucidation of causal relationships between phenotypes have a critical impact on unraveling the mechanisms underlying autoimmune diseases and facilitating the provision of targeted immunotherapies tailored to specific immunophenotypes.

MN, classified as an autoimmune disease, exhibits a strong association with immune cells. However, in current research, distinguishing whether specific immunophenotypic changes are compensatory or causal poses a challenge. One of the focal points of our investigation pertains to the difficulty in discerning the abundance of immune cell subsets that exhibit upstream or downstream relationships with MN. In this study, we have undertaken an unprecedented approach by integrating peripheral blood cell count levels with GWAS data of MN for MR analysis. Our MR analysis has revealed causal associations between specific immune cell subsets in peripheral blood and MN. These cells, having established causal relationships, can act as either upstream or downstream factors in the context of membranous nephropathy. By targeting immune cell subsets associated with upstream factors, potential avenues for immunotherapy can be identified, while those linked to downstream factors hold promise for understanding the development of subsequent conditions following membranous nephropathy. B cells, T cells, autoantibodies, cytokines, and complement system activation collectively contribute to the pathogenesis of MN. An imbalance in the regulatory T cell population is recognized as a prominent characteristic among untreated membranous nephropathy patients(Motavalli et al. 2021). T cells can be divided into three categories according to their activation stages: naïve T cells, effector T cells, and memory T cells. MR analysis results suggest that a decrease in the peripheral blood count of specific CD4+ T cell subsets could potentially increase the risk of MN. In related studies, idiopathic membranous nephropathy patients included in the study exhibited peripheral lymphocytes. Although there were no significant differences in the numbers of peripheral lymphocytes, T cells, and B cells between the two groups, an elevated percentage of CD4+ T cells was observed(Kuroki et al. 2005). Our preliminary inference aligns with these findings, even though the CD4+ T cells in our study were not further divided into T cell subsets.

Studies employing immune cell infiltration analysis have demonstrated significantly higher numbers of CD4 T cells, CD8 T cells, natural killer (NK) cells, monocytes, and macrophages in the renal tissue of membranous nephropathy patients compared to healthy tissue (P<0.001).

Our findings indicate that an increase in the peripheral blood level of CD80 in granulocytes is associated with an elevated risk of developing membranous nephropathy. CD80 plays a role in innate and adaptive immune activation and has been implicated in the pathogenesis of renal disease syndromes. The excessive secretion of cytokines by activated granulocytes can promote inflammation and immune responses, ultimately leading to the deposition of immune complexes in podocytes and resulting in symptoms such as proteinuria.

It is worth noting that CD80 (also known as B7-1) is primarily expressed on antigen-presenting cells and acts as a critical co-stimulatory molecule for T cell activation by binding to the T cell surface receptor CTLA-4. A study involving pediatric and adult patients demonstrated elevated urinary CD80 levels in various glomerular diseases, particularly those with a urinary protein/creatinine ratio ≥3g/g(“Urinary CD80 Discriminates Among Glomerular Disease Types and Reflects Disease Activity - PubMed” n.d.). Interestingly, even in lupus nephritis patients with high levels of proteinuria, urinary CD80 was found to be elevated. However, our research indicates that urinary CD80 is positively correlated with proteinuria and negatively correlated with serum albumin levels, suggesting its potential as a marker for podocyte injury(Zen et al. 2023).

The count of plasmacytoid dendritic cells (pDCs) has also been found to be associated with an increased risk of membranous nephropathy. However, currently, there is a lack of specific research exploring the relationship between membranous nephropathy and dendritic cells.

Conventional dendritic cells type 1 (cDC1) represent a subset of dendritic cells that are primarily involved in cross-presentation to CD8 T cells. Several studies have indicated that the number of cDC1 is associated with the severity of various renal conditions, including acute tubular necrosis, the presence of crescents in immune-mediated glomerulonephritis, interstitial fibrosis in IgA nephropathy and lupus nephritis, as well as the prognosis of IgA nephropathy(Chen et al. 2021). In immune-mediated glomerulonephritis, there is a robust correlation between the number of cDC1 and crescent formation. The abundance of cDC1 in the periglomerular and intraglomerular regions suggests their potential involvement in crescent formation, which may be relevant to some instances of membranous nephropathy associated with crescent formation.

An increase in the count of CD4+ T cells expressing HVEM (Herpesvirus entry mediator) has been shown to be connected to a reduced risk of developing MN. In this context, we would like to introduce the BTLA/HVEM pathway, which is primarily associated with tumor immunity(Yu et al. 2019; “BTLA-HVEM Couple in Health and Diseases: Insights for Immunotherapy in Lung Cancer - PubMed” n.d.). The BTLA (B and T lymphocyte attenuator) molecule acts as a co-inhibitory receptor that inhibits T cell receptor (TCR) signaling in T lymphocytes by recruiting Shp1 and Shp2, which are phosphatases involved in downregulating signaling pathways. Within this pathway, HVEM serves as the ligand for BTLA and can also bind to other molecules such as LIGHT (lymphotoxin-like, exhibits inducible expression, and competes with HSV glycoprotein D for HVEM) or lymphotoxin-alpha (LTα). These interactions can trigger co-stimulatory signals or bind to CD160, another co-inhibitory receptor, at different binding sites. Additionally, HVEM can bind to the glycoprotein D of Herpes Simplex Virus (HSV).

The involvement of the BTLA/HVEM pathway and its relationship with CD4+ T cells expressing HVEM in membranous nephropathy is an area that requires further research and investigation. Please note that the information provided here is a general overview and may not encompass the entire complexity of the BTLA/HVEM pathway or its specific involvement in membranous nephropathy.

An increase in the count of CD4+ T cells expressing HVEM can lead to an increase in the availability of the HVEM ligand. This ligand, in turn, can bind to cytokines unrelated to the pathogenesis of membranous nephropathy and exert effects that antagonize the disease process.

Furthermore, LT-α (TNF-β) and TNF-α are closely related structurally as homologs(Aggarwal, Moffat, and Harkins 1984; Aggarwal, Eessalu, and Hass 1985). TNF-β belongs to the TNF superfamily and shares similarities with TNF-α regarding its ability to activate apoptosis and inflammatory signaling(Etemadi et al. 2013). Similar to TNF-α, TNF-β signals through the lymphotoxin β receptor (LTβR) and can activate both classical and non-classical NF-κB signaling pathways. Additionally, an international genetic study on membranous nephropathy has identified two previously unreported loci, NFKB1 and IRF4, that are associated with inflammation pathways(Xie et al. 2020).

There have been studies demonstrating the association between TNF-α and membranous nephropathy. Firstly, genetic polymorphisms in the TNF-α gene have been found to be associated with membranous nephropathy(Thibaudin et al. 2007). Secondly, serum levels of TNF-α have been observed to be increased in patients with MN and other kidney diseases(Roca et al. 2021).

The MR results indicate that there is no causal association between the peripheral blood count of various CD20-expressing B cell subsets and the risk of developing MN. This finding supports the rationale behind using rituximab (RTX) treatment for MN, as RTX aims to block antibody production by eliminating antibody-producing plasma cells. However, it is essential to note that this treatment approach may face challenges such as drug resistance and varying effectiveness.

Research has indicated that patients with membranous nephropathy have significantly higher counts of plasma cells and regulatory B cells compared to disease-free individuals. Additionally, the expansion of PLA2R-specific memory B cells in vitro has been associated with circulating PLA2R antibody titers(Cantarelli et al. 2020). These findings suggest a compensatory response during the progression of the disease.

Our study possesses several strengths. It represents an innovative endeavor of considerable importance in elucidating the potential influence of genetic susceptibility on changes in peripheral blood cell and immune cell counts in relation to the risk of MN. This aspect remains unexplored in previous MR studies. Our study employed genetic instrumental variables that exhibit strong associations with the exposure. Moreover, these instrumental variables were chosen to be independent of the exposure, outcome, and relevant confounding factors. To mitigate bias in the MR analysis, we adjusted parameters to fulfill the three key assumptions of instrumental variable selection.

However, our study does have certain limitations: In addition to the inherent limitations linked to the requisite assumptions for causal interpretation in MR studies, we did not extensively explore the downstream effects of membranous nephropathy in our research due to the limited advancements in this domain and the unavailability of relevant information. Nonetheless, this underscores a potential avenue for future investigation. Lastly, owing to the limited representation of other ethnicities, the applicability of our study results is restricted to the European population. Future research is required to extend these findings to encompass other ethnic groups.

In conclusion, our study has significantly contributed to advancing our comprehension of membranous nephropathy. Nonetheless, addressing the aforementioned limitations and enhancing the generalizability of these findings to diverse populations necessitate further research endeavors.

## 5 Conclusion

Through MR, we have identified specific immune cell subsets that are associated with membranous nephropathy. These subsets were then analyzed for their upstream and downstream relationships with the disease process. This comprehensive analysis has significantly expanded our understanding of the complex pathophysiology of MN. Furthermore, this new knowledge has opened up potential new avenues for immunotherapy in the treatment of MN.

## 8 Acknowledgments

This is a short text to acknowledge the contributions of specific colleagues, institutions, or agencies that aided the efforts of the authors.

## 9 Data Availability Statement

The datasets (analyzed) for this study can be found in the supplementary document. The datasets (generated) for this study can be found in the address as follow:

Membranous nephropathy data can be obtained from:

https://gwas.mrcieu.ac.uk/datasets/ebi-a-GCST010005/. Immune cell data can be obtained from: https://www.ebi.ac.uk/gwas/publications/32929287. Peripheral blood cell data can be obtained from: https://gwas.mrcieu.ac.uk/datasets/ieu-b-29/ , https://gwas.mrcieu.ac.uk/datasets/ieu-b-30/ , https://gwas.mrcieu.ac.uk/datasets/ieu-b-31/, https://gwas.mrcieu.ac.uk/datasets/ieu-b-32/, https://gwas.mrcieu.ac.uk/datasets/ieu-b-33/, https://gwas.mrcieu.ac.uk/datasets/ieu-b-34/.

